# Changes in central corneal thickness (CCT) and central macular thickness (CMT) following uncomplicated small-incision cataract surgery (SICS)

**DOI:** 10.1101/2020.11.21.20228932

**Authors:** Sanket Parajuli, Ruchi Shrestha, Senny chapagain, Prerana Singh, Ramesh Shrestha

## Abstract

**Objectives:** 1) To study the changes in central corneal thickness (CCT) which is an indirect indicator of corneal endothelial dysfunction after uncomplicated small incision cataract surgery (SICS) and 2) To study changes in macular thickness following uncomplicated SICS.

**Methods:** This was a prospective study conducted in Reiyukai Eiko Masunaga eye hospital, Banepa, Nepal. Those who fulfill inclusion criteria were included in the study. Small incision cataract surgery was performed on 68 eyes of 62 patients Change in central corneal thickness and central macular thickness from baseline was observed post-surgery on 1^st^ day, 1 week and 6 weeks.

**Results:** 33 females and 35 males were included in the study. Mean age was 58.26 years. This difference of visual acuity between pre and post-operative state was statistically significant. The 1^st^ post-operative day (POD) and 1 week POD values when compared with preoperative CCT values were statistically significant. But the 6 weeks POD when compared to preoperative CCT values were not statistically significant. The 1^st^ POD, 1 week POD and 6 weeks POD CMT values when compared with preoperative CMT values were statistically significant.

**Conclusion:** This study revealed that there was a significant rise in CCT after SICS which gradually tended to normalize at 6 weeks. Similarly there was a gradual rise in CMT after SICS persisting even at 6 weeks. However these changes were subtle and there was a marked improvement of visual acuity after SICS.

**Synopsis:** There was statistically significant increase in central corneal thickness and central macular thickness following uncomplicated small incision cataract surgery, the former of which tends to normalize at 6 weeks post-surgery.

## Background

Age related cataract is the most significant cause of bilateral blindness throughout the world. According to World Health Organization (WHO) 47.8% of global blindness is due to cataract. [1] Nepal is one of the first countries in the world where a country-wide survey on blindness was conducted (1980-’81).The prevalence of blindness in 1981AD was found to be 0.84%. [2] The prevalence of blindness reduced from 0.84% in 1981 to an estimated 0.35% in 2011, a reduction of 58%. [3] The reduction in the burden of cataract has obviously been achieved by innumerable cataract surgeries.

Cataract surgery is evolving year after year and we now understand the physiological changes that take place after surgery than ever before. Cataract extraction affect corneal endothelial density. Moderate damage to the endothelium during surgeries can lead to endothelial cell loss and subsequent transient increase in corneal thickness which can be monitored by measurement of central corneal thickness. [4] Researchers have therefore put their effort in the assessment of endothelial damage either by cell counts or by measuring corneal thickness postoperatively. Endothelial cells loss results in an increase in corneal thickness. If severe enough can culminate into corneal decompensation and loss of vision. [5]

Cataract surgery is an invasive procedure that causes inflammatory insult to the eye. [6] Cataract surgery induced surgical trauma resulting in prostaglandins release and blood retinal barriers disruption is thought to be the cause of macular edema. [7] Techniques such as Optical Coherence Tomography (OCT) can help to detect any microscopic changes in the macula. OCT can assess macular thickness quantitatively, it can detect subtle changes of macular thickness and is especially useful to assess the change in macular thickness after cataract surgery at regular intervals. [8]

Taking into consideration the physiological changes that occur after cataract surgery as mentioned above, this study was intended to assess the central corneal thickness (that depicts endothelial cell loss and compensation) and the central macular thickness (macular response to inflammation) after uncomplicated manual small incision cataract surgery (SICS).

## Methods

This was a prospective study study conducted in Reiyukai Eiko Masunaga eye hospital in Banepa, Nepal after ethical approval from local ethical committee. The duration of study was 3 months (1^st^ August 2020 to 30^th^ October 2020). The objectives of the study were 1) To study the changes in central corneal thickness (CCT) which is an indirect indicator of corneal endothelial dysfunction after uncomplicated SICS and 2) To study changes in macular thickness following uncomplicated SICS. Consecutive patients undergoing small incision cataract surgery in the study duration and meeting the inclusion criteria were included in the study. (Figure 1) Written informed consent was taken from all the patients. Sampling techniques was not used since all consecutive patients meeting the criteria were included.

Patients were enrolled in the study if they satisfied the following criteria: 1) Cataract allowing pre-operative CCT (pachymetry) and CMT (OCT) to be performed, 2) patient with good fixation, 3) patients without corneal, retinal or macular pathology, 4) patients who underwent uneventful standard SICS, 5) patient with no glaucomatous changes. Exclusion criteria included 1) mature cataract, 2) traumatic cataract; 3) complicated cataract 4) complication during surgery, 5) corneal, retinal or macular pathology, 6) co-existing glaucoma and uveitis, 7) Astigmatism greater than 2 D 8) Diabetes mellitus.

A complete ocular examination was done for all patients. Examination was done pre-operatively, per-operatively and postoperatively. Pre-operative examination consisted of determination of best corrected visual acuity by Snellen’s chart; Slit lamp examination of the anterior segment; Fundus examination using + 90D lens; CCT (Accutome 4 sight); Central macular thickness using 3D macular scans (Topcon 3D OCT) Per-operative examination consisted of noting the following points: Site of incision, length of incision; total duration of the surgery. Post-operative examination (i.e. follow up on 1 week and 6 weeks) included the following: Determination of the best corrected visual acuity; Slit lamp examination of the anterior segment; fundus examination using +90D lens; and measurements of CCT and CMT. Best corrected visual acuity at 6 weeks was considered as final visual acuity for analysis. Surgical technique was same for all patients. A 6.0 mm PMMA single piece rigid intraocular lens was implanted in the bag at the end of surgery. Any complication during surgery excluded the patients from the study. (Figure 1) Postoperative medications were same for all patients.

Data entry and analysis were done using Statistical Package for Social Sciences SPSS version 25. Comparison was done using the variable means of CCT and CMT using paired t-test and Pearson correlation. The analysis was considered statistically significant association when the P-value was less than 0.05.

## Results

Of 119 eyes which underwent SICS in the study duration, 68 eyes of 62 patients were included in the study. (Figure 1) 33 females and 35 males were included in the study. Mean age was 58.26 years±10 years (Range: 38-80 years) (Table 1)

**Table 1:** Demographic profile of patients

| Variable | Frequency | Percentage |
| --- | --- | --- |
| **Mean age** | 58.26 years ± 10 years | |
| **Gender** |  |  |
| Males: | 35 | 51.4% |
| Females | 33 | 48.6% |
| **Eye undergoing surgery** |  |  |
| Right eye | 26 | 38.2% |
| Left eye | 42 | 61.8% |

Mean duration of surgery was 7.96±2.74 minutes. Mean preoperative visual acuity was 0.95±2.9 log mar unit whereas mean post-operative visual acuity was 0.19±0.21 log mar unit. This difference of visual acuity between pre and post-operative state was statistically significant. (P<0.05) with paired t test at 95% confidence interval. Mean preoperative, 1^st^ post-operative day (POD), 1 week POD and 6 weeks POD CCT values were 528.79±8.49 µm, 575.91±15.65 µm, 546.31±12.7 µm, and 529.97±7.64 µm respectively. (Figure 2) The 1^st^ POD and 1 week POD values when compared with preoperative CCT values were statistically significant with paired t test. (Confidence interval-95%).But the 6 weeks POD when compared to preoperative CCT values were not statistically significant. (Table 2)

**Table 2:** Mean difference in visual acuity, central corneal thickness and central macular thickness in operated eye at various postoperative days in relation preoperative values

| Parameter | value | P* value |
| --- | --- | --- |
| Mean pre Op VA | 0.95 log mar unit |  |
| Mean Post of VA at 6 weeks | 0.19 log mar unit | 0.041 |
| Mean preoperative CCT | $528.79 \pm 8.49 \mu\text{m}$ | .000 |
| Mean CCT - 1<sup>st</sup> POD | $575.91 \pm 15.65 \mu\text{m}$ | .000 |
| Mean CCT - 1 week POD | $546.31 \pm 12.7 \mu\text{m}$ | .000 |
| Mean CCT – 6 weeks POD | $529.97 \pm 7.64 \mu\text{m}$ | .000 |
| Mean CMT -Pre operative | $208.94 \pm 38.78 \mu\text{m}$ | .000 |
| Mean CMT -1<sup>st</sup> POD | $243.90 \pm 36.20 \mu\text{m}$ | .000 |
| Mean CMT- 1week POD | $255.07 \pm 35.16 \mu\text{m}$ | .000 |
| Mean CMT – 6 weeks POD | $246.49 \pm 36.24 \mu\text{m}$ | .000 |

**Notes:** \* paired t test; **Abbreviations:** POD, post-operative day; VA, visual acuity, CCT, central corneal thickness; CMT, central macular thickness

Mean preoperative, 1^st^ POD, 1 week POD and 6 weeks POD CMT values were 208.94±38.78 µm, 243.90±36.20 µm, 255.07±35.16 µm, and 246.49±36.24 µm. (Figure 3) The 1^st^ POD, 1 week POD and 6 weeks POD CMT values when compared with preoperative CMT values were statistically significant with paired t test. (Confidence interval-95%). (Table 2) Bivariate analysis (Pearson) of duration of surgery and CCT at 1^st^ POD did not show a significant correlation. Similarly bivariate analysis (Pearson) of duration of surgery and CMT at 1^st^ POD did not show a significant correlation.

## Discussion

Mean age of patients in our study population was 58.26 years. The presence of elderly population in our study group reestablishes the aging process as the commonest etiology of uncomplicated cataract. Males constituted a larger proportion in the study. This was similar to study conducted by Mganaga et al where cataract was seen mostly in the elderly population and in males. [9]

Endothelial cell loss, which clinically presents as corneal edema, acts as a measure for estimating the safety of the surgical technique. Our study showed that in manual small incision cataract surgery, the mean CCT on day 1st postoperative increased from 528.79 µm baseline CCT to 575.91 µm. Mean CCT was 546.31±12.7 µm on 1 week POD and slight decrease to 529.97 was noted on 6 week POD. Hence, it shows that there was some endothelial cell loss leading to a change in corneal thickness in first few weeks of surgery but was well compensated at 6 weeks POD. This is similar to study conducted by Jacob that reported that maximal increase occurred within 24 hours following cataract surgery. [10] This study was similar to studies conducted by Bolz et al, Ventura et al and Cheng et al where increase followed by gradual decreases in corneal thickness was recorded following cataract surgery. [11-13]

Subclinical changes may occur in macular thickness without the visual acuity being affected. In this study there was a statistically significant increase in post-operative CMT values from baseline. Gharbiya et al demonstrated a progressive increase in retinal thickness after uncomplicated cataract surgery. Only at 6 months after surgery, retinal thickness tended to normalize in the central fovea. [14] Yoo et-al studied the changes in central subfield macular thickness (CSMT) using OCT after cataract surgery. They concluded at 1 month CSMT significantly increased by 22.2 ± 47.10 µm after cataract surgery in the operated eye compared with the fellow eye (p = 0.01). [15] There was a significant improvement in BCVA. The improvement in vision was statistically significant (P value 0.041). It, thus, concludes that there is endothelial cell loss leading to increased CCT values and an increase in central macular thickness but not to the extent to cause visual impairment following uncomplicated SICS in healthy individuals.

One of the limitations of our study is that only one technique of SICS is used. Other techniques may give different results. Duration of study could have been much longer. CCT is only an alternative for endothelial cell function. Ultrasound pachymetry was used instead of specular microscopy to assess endothelium function. Also, measurements estimated by ultrasonic pachymetry are more variable. The patients were from a specific region of the country belonging to certain ethnic groups only and thus the results cannot be generalized.

## Conclusion

This study revealed that there was a significant rise in CCT after SICS which gradually tended to normalize at 6 weeks. Similarly there was a gradual rise in CMT after SICS persisting even at 6 weeks. However these changes were subtle and there was a marked improvement of visual acuity after SICS.

## Supporting information

strobe checklist

## Data Availability

The authors confirm that the data supporting the findings of this study are available within the article. Data can also be provided on request by the corresponding author

## Abbreviations

(CCT): Central Corneal Thickness
(CMT): Central macular thickness
(SICS): Small-Incision Cataract Surgery
(POD): Post-operative day
(CSMT): Central subfield macular thickness
(OCT): Ocular coherence tomography

## Declarations

- Ethics approval and consent to participate: Approved by local ethical committee of Reiyukai Eiko Masunaga eye hospital (ERB-REH 20/04), written informed consent from all participants were obtained. All applicable international, national, and institutional guidelines were followed.
- Consent for publication: provided to the journal
- Competing interests: None
- Conflict of interests: None
- Funding: None
- Availability of data: The authors confirm that the data supporting the findings of this study are available within the article. Data can also be provided on request by the corresponding author.
- Author contributions: SP drafted the manuscript. SC, RS made the plan of study and made the proforma. PS and RS collected the data and made sure patient were on regular follow up visits.
- Acknowledgement: N/A

