## Supplementary material for "Changes in central corneal thickness (CCT) and central macular thickness (CMT) following uncomplicated small-incision cataract surgery (SICS)": figure

Figure 1: Patient inclusion and exclusion

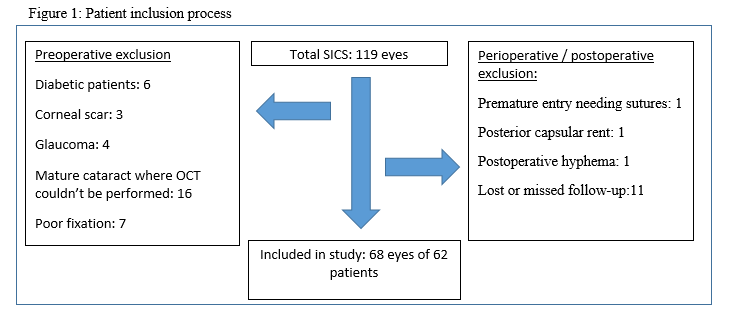

CCT

|  | CCT |
| --- | --- |
| preoperative | 528.79 |
| 1st POD | 575.91 |
| 1 week POD | 546.31 |
| 6 weeks POD | 529.97 |

Figure 2: Progression of mean central corneal thickness at different days of surgery

CMT

|  | CMT |
| --- | --- |
| Preoperative | 208.94 |
| 1st POD | 243.9 |
| 1 week POD | 243.9 |
| 6 weeks POD | 243.9 |

Figure 3: Progression of mean central macular thickness at different days of surgery
